# Factors impacting e-learning in health sciences education: a protocol for systematic review and meta-analysis

**DOI:** 10.1101/2020.06.26.20140566

**Authors:** Krishna Regmi, Linda Jones

## Abstract

E-learning has been widely used in higher education as it provides better access to learning resources online, utilising technology – regardless of learners’ geographical locations and timescale – to enhance learning. Despite growing evidence claiming that e-learning is as effective as traditional means of learning, there is very limited evidence. This protocol aims to assess the effects of e-learning factors that impact on health sciences education (HSE). We will conduct a systematic review meta-analysis research of both randomised controlled trials and non-randomised controlled trials. Seven databases – MEDLINE, EMBASE, Allied & Complementary Medicine, DH-DATA, PsycINFO, CINAHL, and Global Health – will be searched, from 1980 through 2020. The PRISMA-P checklist will be used while preparing this protocol. Two reviewers will independently screen the papers and extract data. We will utilise the JBI quality appraisal tools for quantitative and qualitative studies, and Mixed Methods Appraisal Tool appraisal tools to assess the quality. A narrative synthesis, using thematic analysis, will be conducted for the included studies. If sufficient data are available, the random-effects model for meta-analysis will be performed to measure the effect size of e-learning on health sciences education or the strengths of relationships. The outcome of this review will provide a useful checklist of potential factors to develop an e-learning approach in HSE. This might provide a basis for developing the best methods of e-learning in education so that e-learning policy in education and learning settings in the HSE context could be administered effectively, efficiently and equitably.

## Introduction

There are different meanings or interpretations of e-learning, but employing the technology to provide online access to learning resources for the improvement of learning is the principal aspect of e-learning (Holmes & Garder, n.d.; Sandars, 2013). E-learning has been defined as an educational method that would facilitate learning by the application of information technology and communication (Golband et al., 2014). Recently, e-learning has been well recognised in medical education and other healthcare disciplines that included the dental, public health, nursing, and other allied healthcare education, which this protocol refers to as health sciences education (HSE). However, the impact of e-learning and its effect on learners’ performance or enhancing their learning has been well debated.

Similarly, e-learning has had less impact than intended, and HSE practices have remained largely unchanged over the past decade. Cook et al. (2008) raise some concerns over whether e-learning would actually enhance learning, particularly ‘the extent to which knowledge-based learning compared with alternative approaches to medical education’. Though some literatures on e-learning have provided some promises that e-learning would be equally effective as traditional methods of learning or teaching, still there is very limited evidence demonstrating when and how best e-learning enhances education and learning, and the factors associated with it (Childs et al., 2005; Cook et al., 2008; Curran & Fleet, 2005; Donnelly et al., 2012; McCutcheon et al., 2015; Wutoh et al., 2004). As Kim (2006) argues, most of the published evidences appear to have three major limitations: (a) they are mostly descriptive, (b) they have clearly failed to demonstrate the outcome measures, and (c) the majority have faults due to weakness or inappropriateness in study designs.

Another systematic review, capturing 176 empirical studies, conducted between 1996 and 2008, shows that students in online conditions performed modestly better as compared to those learning the same material through traditional face-to-face instruction (Means et al., 2010). These interpretations, however, should be treated with caution, as the conditions and dimensions for both methods are not the same, particularly the learners’ and facilitators’ time spent on setting or accomplishing tasks, level of accessibility, and convenience (Cook et al., 2010). A Cochrane Review involving 5679 health professionals, published in 2018, examining the effects of e-learning versus traditional learning, reported little or no differences in patient outcomes or health professionals’ skills and behaviours (Vaona et al., 2018).

Similarly, several studies make claims for e-learning and learning enhancement, but the results appeared rather mixed (Cappel & Hayen, 2004; Cook et al., 2008; Ruiz et al., 2006). It has been found that if we simply compare the outcomes between e-learning and no training interventions, e-learning is generally far more effective in gaining knowledge and skills including positive behaviours, but this does not necessarily mean that the results are significant mainly due to the fact that results are heterogeneous (i.e. inconsistent results) and are frequently in small studies (Al-Shorbaji et al., 2015; Fletcher, 2007). Until now there has been limited, scattered and patchy evidence in relation to the effects of e-learning for HSE (Al-Shorbaji et al., 2015; Kim, 2006). Our preliminary scan showed that no earlier systematic review and meta-analysis protocol has addressed the objectives of this systematic review.

## Aims and objectives

The aim of this review protocol is to assess the effects of e-learning factors that impact on health sciences education.

### Primary objective

The primary objective is to identify, appraise and synthesise the existing evidence on barriers and facilitators to e-learning in HSE.

### Secondary objective

A secondary objective is to quantify and analyse to measure the effect of e-learning on health sciences education or the strengths of relationships.

To the best of our knowledge, this is the first systematic review and meta-analysis research protocol examining and synthesising the factors – enablers or barriers – evidencing to e-learning in making health sciences education effective.

## Methods

### Study design

This study will utilise a systematic review and meta-analysis method, which will consider both randomised controlled trials and non-randomised trials (prospective and retrospective observational studies) of good-quality studies. Chalmers and Atlman (1995) define systematic review as a “review that has been prepared using a systematic approach to minimise biases and random errors which is documented in a materials and methods section.” Meta-analysis includes the statistical analysis for combining the results of a number of individual studies to produce summary results, e.g. pooled research studies (Khan et al., 2011). The Preferred Reporting Items for Systematic Reviews and Meta-Analysis Protocols (PRISMA-P) checklist has been used in the preparation of this protocol (Shamseer et al., 2015).

### Search strategies

A broad search strategy will be designed to maximise the level of sensitivity to identify potential studies and specificity to identify definitely relevant studies in searching (Higgins et al., 2019), and improve both the recall and precision ratios (Katcher, 2006). A systematic structured literature search of seven databases – MEDLINE, EMBASE, Allied & Complementary Medicine, DH-DATA, PsycINFO, CINAHL, and Global Health – will be conducted, from 1980 through 2020. Primary search terms are e-learning (all synonyms) and health sciences education (all synonyms) using ‘Textword searching’ – searching for a word or phrase appearing anywhere in the document, where the document is the citation (article title, journal name, author), not the full text of an article, and ‘Thesaurus (MeSH, EMTREE) searching’, employing Boolean operators and truncations (Table 1).

**Table 1.**
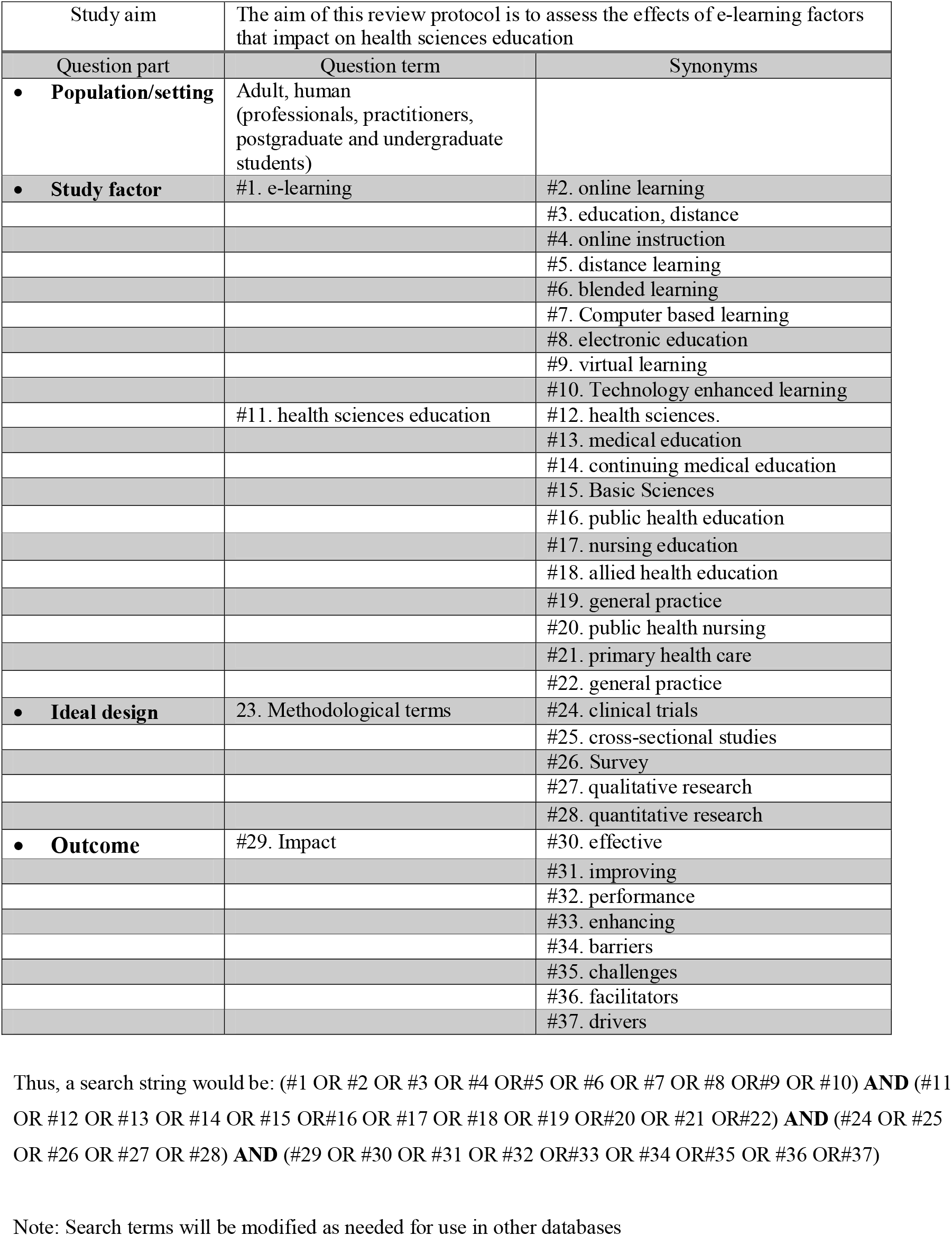
Search strategy for the MEDLINE

|  |  |  |
| --- | --- | --- |
| Study aim | The aim of this review protocol is to assess the effects of e-learning factors that impact on health sciences education |  |
| Question part | Question term | Synonyms |
| • **Population/setting** | Adult, human (professionals, practitioners, postgraduate and undergraduate students) |  |
| • **Study factor** | #1. e-learning | #2. online learning |
|  |  | #3. education, distance |
|  |  | #4. online instruction |
|  |  | #5. distance learning |
|  |  | #6. blended learning |
|  |  | #7. Computer based learning |
|  |  | #8. electronic education |
|  |  | #9. virtual learning |
|  |  | #10. Technology enhanced learning |
|  | #11. health sciences education | #12. health sciences. |
|  |  | #13. medical education |
|  |  | #14. continuing medical education |
|  |  | #15. Basic Sciences |
|  |  | #16. public health education |
|  |  | #17. nursing education |
|  |  | #18. allied health education |
|  |  | #19. general practice |
|  |  | #20. public health nursing |
|  |  | #21. primary health care |
|  |  | #22. general practice |
| • **Ideal design** | 23. Methodological terms | #24. clinical trials |
|  |  | #25. cross-sectional studies |
|  |  | #26. Survey |
|  |  | #27. qualitative research |
|  |  | #28. quantitative research |
| • **Outcome** | #29. Impact | #30. effective |
|  |  | #31. improving |
|  |  | #32. performance |
|  |  | #33. enhancing |
|  |  | #34. barriers |
|  |  | #35. challenges |
|  |  | #36. facilitators |
|  |  | #37. drivers |

Thus, a search string would be: (#1 OR #2 OR #3 OR #4 OR#5 OR #6 OR #7 OR #8 OR#9 OR #10) **AND** (#11 OR #12 OR #13 OR #14 OR #15 OR#16 OR #17 OR #18 OR #19 OR#20 OR #21 OR#22) **AND** (#24 OR #25 OR #26 OR #27 OR #28) **AND** (#29 OR #30 OR #31 OR #32 OR#33 OR #34 OR#35 OR #36 OR#37)
Note: Search terms will be modified as needed for use in other databases

The search structure will consist of the following:

1. Break down the search into separate concepts.
2. Search each concept separately using free and MeSH terms to obtain four sets of results.
3. Combine the four sets so that both concepts will be in the same references.
4. Consider using relevant limits (e.g. English, humans, age groups).

The ‘Related Articles’ feature in PubMed will be consulted. Searches will also be supplemented by reviewing the reference lists (‘references of references’) of selected articles to find any other relevant papers. We will also ask subject experts/information specialists from both Universities (Bedfordshire and Dundee) to verify the research strategy, ensuring its comprehensiveness.

### Inclusion and exclusion criteria

It has been argued that inclusion and exclusion criteria for any research should be set up in line with the proposed aim and objectives, ensuring that peer-review articles will not only be drawn within the context and boundaries of the research objectives, but also will be structured in a way that would enable the researcher to answer the proposed study objectives (Centre for Reviews and Dissemination, 2008). The inclusion and exclusion criteria for this study have been provided.

### Criteria for considering studies for review

#### Inclusion criteria

- Type of studies: Any primary study designs, both randomised controlled trials and non-randomised trials (prospective and retrospective observational studies) that reported or identified or quantified barriers and facilitators to e-learning will be eligible.
- Journal quality: Articles published in peer-reviewed journals.
- Articles discussing factors – barriers or facilitators – about el-HSE as outcome measures.
- Language and time limit: Articles published in English language and published after 1980.

#### Exclusion criteria

- Articles published in secondary, non-empirical studies or grey literature.
- Commentaries, review documents, case studies, letters, discussion papers, posters, conference abstracts, congress reports and dissertations.
- Articles not published in peer-reviewed journals.
- Articles falling outside el-HSE.
- No full text available.
- Articles not published in English language and published before 1980.

### Participants

Enablers or barriers related to e-learning would vary across different institutions and stakeholders. In order to capture the broad-spectrum eligible population, we will consider professionals, practitioners, postgraduate and undergraduate students as well as policy-planners, policy-makers or researchers, and administrators including continuing professional development for healthcare professionals within the fields of medical, dental, public health, nursing, and other allied healthcare educations.

### Interventions

Any interventions i.e. computer technologies used to facilitate the provision of online access to learning resources for the improvement of learning would be considered, e.g. e-learning, online learning, online instruction, distance learning, distance teaching or computer-assisted instruction.

### Control/comparators

Studies will be included regardless of control/comparator group.

### Study outcomes

For the purpose of this study, any type of barriers or enablers to e-learning, primarily in making learning effective, improving learning performance, or enhancing learners’ level of knowledge, skills and attitudes would be considered as outcomes. The conceptual meaning of barriers and facilitators has been borrowed from Reynders et al. (2016), i.e. barriers refer to any variable that impedes or obstructs, whereas facilitators would act other way round, e.g. variables that would ease and/or promote the use of e-learning in HSE.

### Measure of effect

For dichotomous data, we will measure the risk ratios (RR). Mean differences, and where appropriate, standardised mean differences will be used for continuous data.

### Screening and data extraction

The literature which will emerge from the databases, snowballing and hand-searching, will be screened at two stages: first, a review of abstracts and titles of the retrieved literature to see whether they will meet minimum inclusion criteria, i.e. whether the paper will address el-HSE as this review defines it, and whether the paper will report data on factors – enablers and challenges related to learning performance or outcomes, and articles will be removed which do not meet a minimum criterion at this stage. Screening of titles and abstracts will be done with two reviewers. Second, the full text of the included articles will be reviewed using the JBI Critical Appraisal Checklists for qualitative, quantitative studies (Lockwood et al., 2015; Munn et al., 2015) and Mixed Methods Appraisal Tool (MMAT) for mixed-method studies (Hong et al., 2018). At this stage, more articles might be excluded. As Means et al. (2010) argue, the intent of the two-stage approach is to gain efficiency without risking exclusion of potentially relevant, high-quality studies of online learning effects. The standard PRISMA flow diagram (Fig. 1) will be used to provide the process of study selection (Moher et al., 2009).

**Figure 1.**
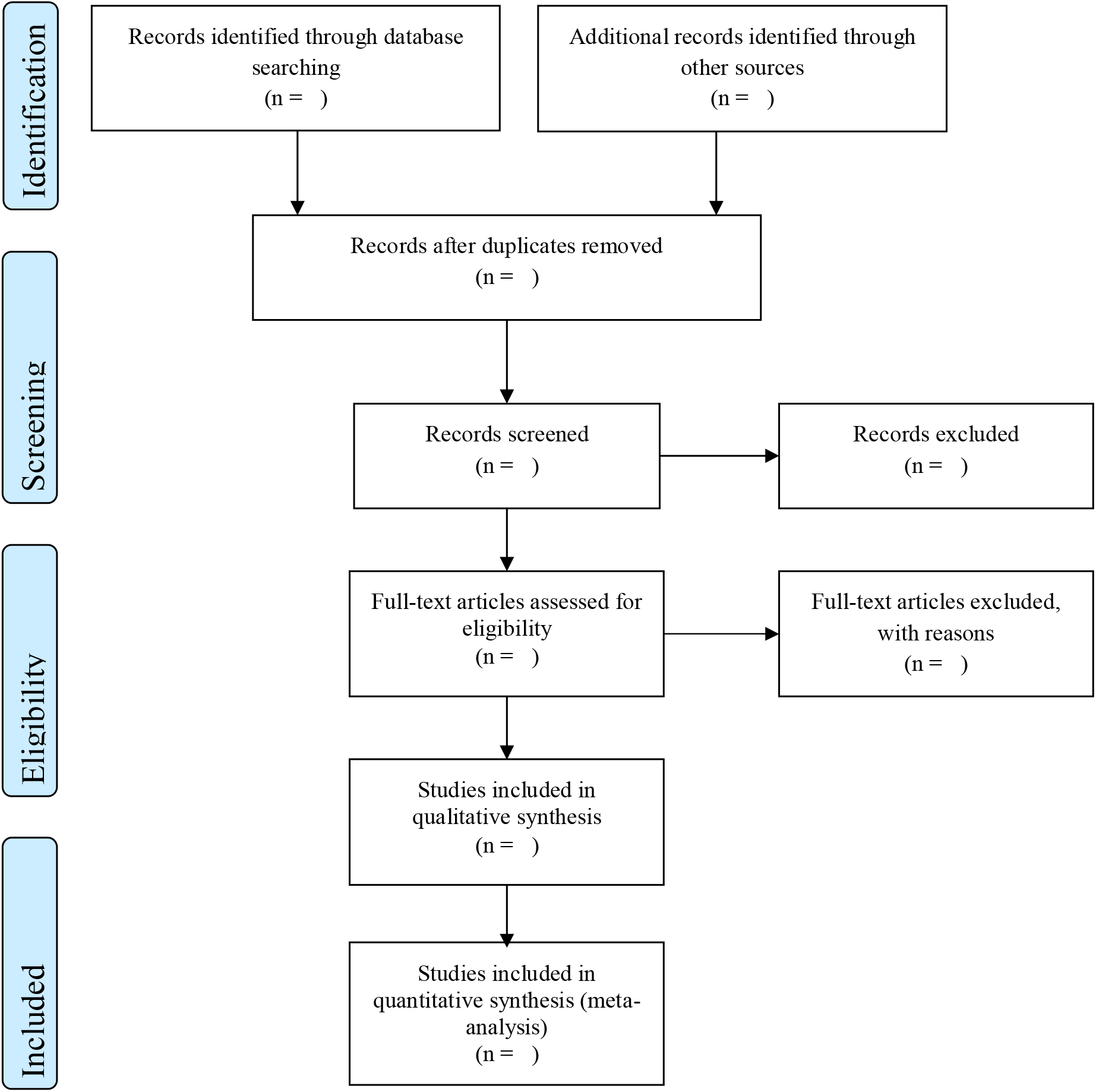
PRISMA flow diagram.

### Quality assessment and risk of bias

We will utilise the quality assessments using critical appraisal tools (Lockwood et al., 2015; Munn et al., 2015; Hong et al., 2018) to assess the methodological qualities including the possibility of bias in study design, conduct and analysis. The results of these appraisals will be used to inform the synthesis and interpretation of the study results. Both JBI tools and MMAT have established content validity and have been piloted across all methodologies (Kerins et al., 2018; Pluye, 2015; Souto et al., 2015). The retrieved papers will be assessed by two reviewers (KR and LJ) using the standardized 10-item, 9-item and 5-item critical appraisal checklists for qualitative assessment, quantitative and mixed methods studies, respectively (Table 2).

**Table 2.** Results from the critical appraisal of methodological quality

| Results from critical appraisal of qualitative studies - JBI Critical Appraisal Checklist for Qualitative Research (Lockwood et al., 2015) |  |  |  |  |  |  |  |  |  |  |
| --- | --- | --- | --- | --- | --- | --- | --- | --- | --- | --- |
| Studies no/Question no | 1 | 2 | 3 | 4 | 5 | 6 | 7 | 8 | 9 | 10 |
| In total |  |  |  |  |  |  |  |  |  |  |
| Results from critical appraisal of quantitative study - JBI Critical Appraisal Checklist for Quantitative Research or Prevalence Data (Munn et al., 2015) |  |  |  |  |  |  |  |  |  |  |
| Study no/question no | 1 | 2 | 3 | 4 | 5 | 6 | 7 | 8 | 9 |  |
| In total |  |  |  |  |  |  |  |  |  |  |
| Results from critical appraisal of mixed methods study - Mixed Methods Appraisal Tool (Hong et al., 2018) |  |  |  |  |  |  |  |  |  |  |
| Study no/question no |  |  |  |  |  |  |  |  |  |  |
| Qualitative components | 1.1 | 1.2 | 1.3 | 1.4 | 1.5 |  |  |  |  |  |
| Quantitative components | 4.1 | 4.2 | 4.3 | 4.4 | 4.5 |  |  |  |  |  |
| Mixed methods | 5.1 | 5.2 | 5.3 | 5.4 | 5.5 |  |  |  |  |  |
| In total |  |  |  |  |  |  |  |  |  |  |

To facilitate comparison of appraisal processes, both reviewers will record the rationale for inclusion or exclusion, and discrepancies will be discussed and resolved by consensus and, in the event a consensus cannot be reached, a third reviewer will arbitrate.

### Assessment of reporting biases

Publication bias, often called reporting bias and dissemination bias, refers to the concern that studies which report relatively large effects are more likely to be published as compared to studies reporting smaller effects (Borenstein, 2019, p.155). Similarly, published studies that include multiple outcomes would be more likely to report the outcomes than if they showed statistically significant results (Sterne et al., 2008). One approach to address the publication bias is to follow the Trim and Fill procedures, i.e. assessing asymmetry or symmetry in the Funnel plot if more than 10 eligible studies are identified. This approach would estimate the extent of bias or estimate of the adjusted effect size (Duval & Tweedie, 2000). We will use this approach while assessing the publication bias in the included studies, but Borenstein (2019, p.165) warns that the presence of bias will not automatically invalidate the results.

### Data analysis and synthesis

A narrative synthesis, using thematic analysis, will be conducted for the included studies. Both qualitative and quantitative approaches will be used for data analysis. We will also provide a descriptive numerical summary. Based on the nature of the proposed study topic, we anticipate that included studies would be heterogeneous which preclude meta-analysis.

If sufficient data are available, i.e. identical on important factors and addressing the same fundamental question, to make an inference to a universe of comparable studies, the random-effects model for meta-analysis will be employed for the analysis to measure the effect size of e-learning on health sciences education or the strengths of relationships using the software Comprehensive Meta-Analysis (CMA, version 3. https://www.meta-analysis.com/pages/new_v3.php?cart=BT2P4569026). The purpose of using a random-effects model in the analysis is “to incorporate the assumption that the different studies are estimating different, yet related, intervention effects” (Higgins et al., 2019). To assess the heterogeneity of effects, I^2^ together with the observed effects (Q-value, with degrees of freedom) will be used to provide the true effects in the analysis. Q-value is the sum of the squared deviations of all effect sizes from the mean effect size. Generally, this value is on a standardised scale, so that a large deviation gets more weight if the estimate is precise, and less weight if the estimate is imprecise (Borenstein et al., 2009). In fact, I^2^ statistics does not tell us how much heterogeneity there is, but it tells what proportion of the observed variance reflects in true effect sizes rather than the sampling error. As such, it provides some context for understanding the forest plot (Borenstein et al., 2017). If I^2^ statistics is low (near zero), then most of the variable in the forest plot is due to sampling error. Conversely, if I^2^ statistics is very high (say, more than 75%) then most of the variance in the forest plot is due to variance in true effects. If we could somehow plot the variance of true effects, most of the variance would remain (Higgins et al., 2019).

### Tabulating the included studies

Data from eligible studies will be extracted independently by two reviewers based on the guidelines produced by the Cochrane Group (https://ph.cochrane.org/review-authors) (Table 3). As Rodgers and colleagues confirm, this would not only improve the process of transparency by better understanding what sorts of data extracted from which studies, but also recognising the contribution made by each study to the overall synthesis (Rodgers et al., 2009). In addition, such tables will demonstrate how the individual study area contributes to the reviewers’ final conclusion.

**Table 3.**
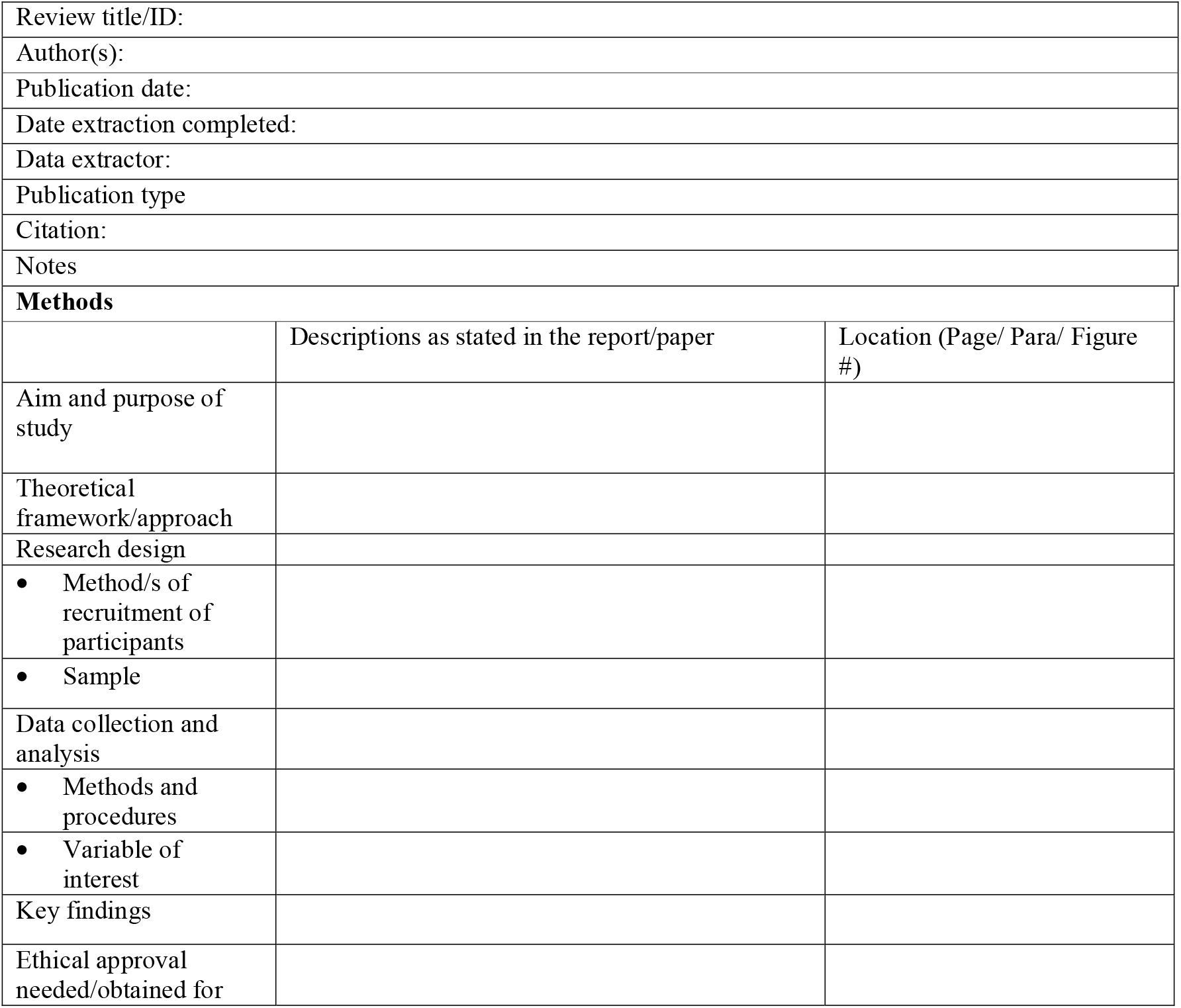
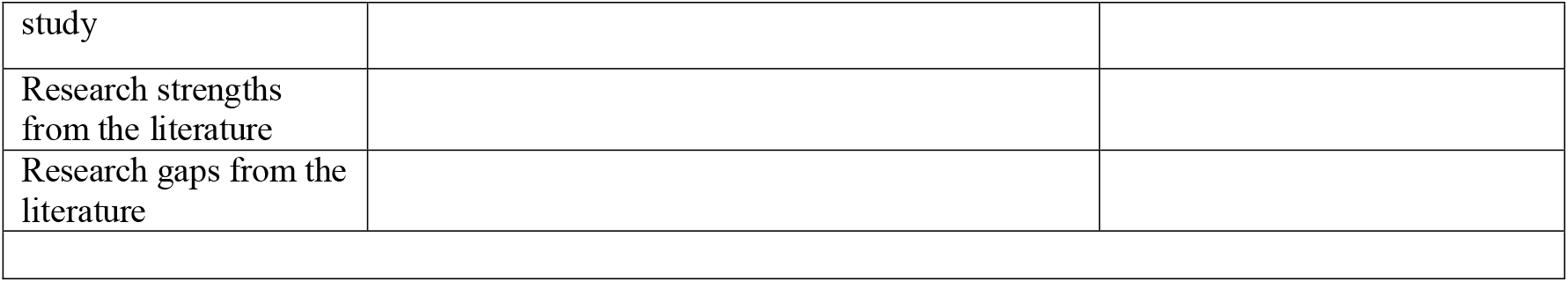
Data Extraction Template

|  |  |  |
| --- | --- | --- |
| Review title/ID: |  |  |
| Author(s): |  |  |
| Publication date: |  |  |
| Date extraction completed: |  |  |
| Data extractor: |  |  |
| Publication type |  |  |
| Citation: |  |  |
| Notes |  |  |
| **Methods** |  |  |
|  | Descriptions as stated in the report/paper | Location (Page/ Para/ Figure #) |
| Aim and purpose of study |  |  |
| Theoretical framework/approach |  |  |
| Research design |  |  |
| • Method/s of recruitment of participants |  |  |
| • Sample |  |  |
| Data collection and analysis |  |  |
| • Methods and procedures |  |  |
| • Variable of interest |  |  |
| Key findings |  |  |
| Ethical approval needed/obtained for |  |  |

|  |
| --- |
| study |
| Research strengths from the literature |
| Research gaps from the literature |

### Dealing with missing data

In the case of missing data that might be important to summarise/synthesise the findings of the study or details of the studies are unclear, corresponding authors of included studies will be contacted.

### Sub-group analysis

We will group by the types of e-learning (e.g. enhanced or adjunct learning – face-to-face learning, blended e-learning model – face-to-face and online learning, and pure online or fully-online learning), by respondents (e.g. medical, nursing and allied healthcare professionals or practitioners), by student (post-graduate and undergraduate).

### Risk of bias assessment

Risk of bias will be examined, as it provides the variation, e.g. heterogeneity in the results of the studies included in the study. As Higgins et al. (2019) argue, rigorously conducted studies in the systematic review would provide more truthful results, and the results from the studies of variable validity would give either false negative or false positive conclusions. Therefore, assessing the risk of bias in all studies in any review is important. In assessing risk, we will create a table with a row for every relevant type of potential bias, and then classify each study on each row as having a low, unclear, or high risk of bias. In this study, the issue of bias will be kept separate from the core analysis – meaning analysis will be performed without worrying about the quality/bias. We will then use the risk of bias table to provide the context for the analysis (Borenstein, 2019). As Borenstein (2019) suggests, “if the analysis shows a clinically and/or substantially important effect, we will assess the entirety of the evidence by considering the risk of bias as well” (p.326). Generally, the bias table provides the type of bias (e.g., selective reporting of outcomes, random sequence generation, allocation of concealment, blinding of participants, personnel and assessors, incomplete outcome data and other potential threats to validity) in each study. If, for example, most rows are unshaded then that it is considered a low risk of bias, whereas if some (or all) rows are either partly shaded or dark (risk of bias will be either unclear or high), this would provide relatively less confidence in the results (Higgins et al., 2019). Generally, risk of bias assessment tool will depend on the study type. We will use RoB 2 tool (Sterne et al., 2019) for randomised and ROBINS-I tool (Sterne et al., 2016) for non-randomised trials while assessing the risk of bias.

### Ethics

As this is a protocol for a systematic review and meta-analysis, neither patients nor public participation will be directly involved, and ethics approval and consent will not be required either.

### Dissemination

We will be able to disseminate the study findings using the following strategies: first, a copy of the final systematic review and meta-analysis protocol will be made available at the libraries of both authors’ universities (Dundee and Bedfordshire), so that students and staff will be able to gain benefits, largely on how to write a systematic review protocol. Second, we will be published in an academic peer-reviewed journal. Third, an abstract will be presented at suitable national/international medical education or e-learning conferences or workshops.

## Discussion

To the best of our knowledge, this will be the first systematic review and meta-analysis protocol to identify and quantify the potential factors (barriers and facilitators), and analyse to measure the effect size of e-learning on health sciences education or the strengths of relationships. Some evidence suggests that e-learning facilitates the process of learning and thereby changes in practice by supporting instructional design and delivery mechanisms, which captures the developing of materials using set learning objectives, including teaching strategies – embedding feedback and evaluation to influence learners’ intrinsic and extrinsic motivation factors. However, how these factors, in fact, would influence examining and synthesising internal, external and contextual factors has not been well researched in the past (Lewis et al., 2014). Second, there are some frameworks and models of online education which employ different methods and tools to collect or gather data, and they mostly used pre- and post-test questionnaires or self-assessment checklists (Kirkpatrick, 1998; Levy, 2006; Phillips, 2000; Stufflebeam, 2001). Similarly, another model was more focused on motivating learners and e-moderators in general education, but none of them is specifically designed or developed in line with medical or health sciences education (HSE) (Mayes, 2004).

These descriptions have illustrated two points: first, a lot of health sciences (or professional) education e-learning research and evaluation clearly lacks some theoretical perspectives. Second, most of the conceptual and theoretical work is based on neither medical nor health sciences education, but several pieces of work look at education generally.

The potential limitations of this study would be that if the retrieved studies would be heterogeneous in nature, we might not be able to perform a meta-analysis measuring the effect size of e-learning on health sciences education or the strengths of relationships. Second, there will be a small number of suitable articles to identify, appraise and synthesise the existing evidence on barriers and facilitators related to e-learning with different disciplines, i.e. nursing, physiotherapy, public health, allied healthcare sciences, etc. Third, this research is unfunded, and both time and resource might be limited. Therefore, articles published in English only will be included in this study.

In light of the identified limitations or challenges, one of the major strengths of this study is to apply transparent methods and approaches for systematic review and meta-analysis so that future researchers, practitioners and academics will be able to reproduce its methodology easily. Similarly, the outcome of this review will provide a useful checklist of potential factors to develop an e-learning approach in HSE. This might provide a basis for developing the best methods of e-learning in education so that e-learning policy in education and learning settings in the HSE context could be administered effectively, efficiently and equitably.

## Data Availability

Not applicable

## Acknowledgements

This research protocol is based on ongoing research studies undertaken for medical education by KR at the University of Dundee, UK. The qualitative component of the study is published elsewhere (Regmi & Jones, 2020).

